# A snapshot of a pandemic: the interplay between social isolation and COVID-19 dynamics in Brazil

**DOI:** 10.1101/2021.04.29.21256267

**Authors:** Cláudia P. Ferreira, Diego Marcondes, Mariana P. Melo, Sérgio M. Oliva, Cláudia M. Peixoto, Pedro S. Peixoto

## Abstract

In response to the COVID-19 pandemic, most governments around the world implemented some kind of social distancing policy in an attempt to block the spreading of the virus within a territory. In Brazil, this mitigation strategy was first implemented in March 2020 and mainly monitored by social isolation indicators built from mobile geolocation data. While it is well known that social isolation has been playing a crucial role in epidemic control, the precise connections between mobility data indicators and epidemic dynamic parameters have a complex interdependence. In this work, we investigate this dependence for several Brazilian cities, looking also at socioeconomic and demographic factors that influence it. As expected, the increase in the social isolation indicator was shown to be related to the decrease in the speed of transmission of the disease, but the relation was shown to depend on the urban hierarchy level of the city, the human development index and also the epidemic curve stage. Moreover, a high social isolation at the beginning of the epidemic relates to a strong positive impact on flattening the epidemic curve, while less efficacy of this mitigation strategy was observed when it has been implemented later. Mobility data plays an important role in epidemiological modeling and decision-making, however, we discuss in this work how a direct relationship between social isolation data and COVID-19 data is hard to be established. Understanding this interplay is a key factor to better modeling, for which we hope this study contributes.

## 1 Introduction

The importance of human mobility during the COVID-19 outbreak has been clear since the beginning of the epidemic, in Wuhan, China in late December 2019, from where it spread throughout the highly connected network of tourism and business cities in the world, becoming a pandemic on March 11, 2020. It is not the first disease that used this pathway to move, since SARS and H1N1, for example, have also used it, but is certainly the most efficient one to do so in the last century. Since it is transmitted mainly through contaminated droplets of naso-oropharyngeal secretions from infected individuals, the rate of infections by the SARS-Cov-2 is related to human mobility in general, but more specifically to the close contacts between individuals of a population^23^.

As an emerging disease, very little was known about it, and, until the development of vaccines, there was no definitive scientific-based medical treatment or pharmaceutical prevention method for it. Therefore, its control was based mainly on non-pharmaceutical methods seeking either to reduce the odds of contact with an infected individual causing an infection, such as mask-use and hand washing, or to avoid the contact between an infected and susceptible individual, such as social distancing, lockdown, and detection and isolation of infected individuals. While the efficacy of mask-use and hand-washing, for example, may be quantified under controlled circumstances^37^, the efficacy of measures aiming to diminish the contacts between individuals through the decrease of human mobility may be hard to quantify. Besides, human mobility measures such as isolation indexes are often not accurate or hard to interpret^31^. In summary, the spread of COVID-19 is a multi-faceted complex process that is influenced by many uncontrollable variables, which may mask the effect of human mobility on it^8^, so it is difficult to establish a cause and effect relationship between an index of human mobility and the rate of infections^20^.

Nevertheless, the pandemic spread has been frequently modeled by dynamical systems (deterministic or stochastic models)^18^ which usually have as control parameter some measure of amplification/attenuation of the disease infection rate due to different transmission scenarios of varying human contact. Human contact is indirectly measured through indices of human density, mobility, isolation, and social distancing. While in theory this is well accepted and provides insights on possible outcomes of the pandemic, it is usually very difficult to quantify the effects of non-pharmaceutical interventions on infection rates^3,7,9^, so most studies use *ad hoc* tuning parameters^40,38^.

Focusing on Brazil, it is the largest and most populated country of South America with around 210 million inhabitants. Although it is the 9th world economy, according to the International Monetary Fund^27^, it is a country marked by great inequalities and underdevelopment. Brazil is divided into five disparate regions (see Figure 1). The South and Southeast regions concentrate more than half the population, have the most developed infrastructure, are home to the wealthiest states and financial centers, offer better-paying jobs, and have better socio-economic indexes. The Midwest region, the least populated of them, is home to the country capital city Brasília, has a lower population density, consisting most of the agricultural land, and has socio-economic indexes lower than the southern regions, with the exception of the district of the capital city, which has better indexes. The North and Northeast regions are the poorest, with lower socio-economic indexes and underdeveloped infrastructure. Although both regions suffer from the lack of infrastructure, the situation is worse in the North, whose territory is covered by the Amazon rain forest, which makes logistics difficult in the region.

**Fig. 1.**
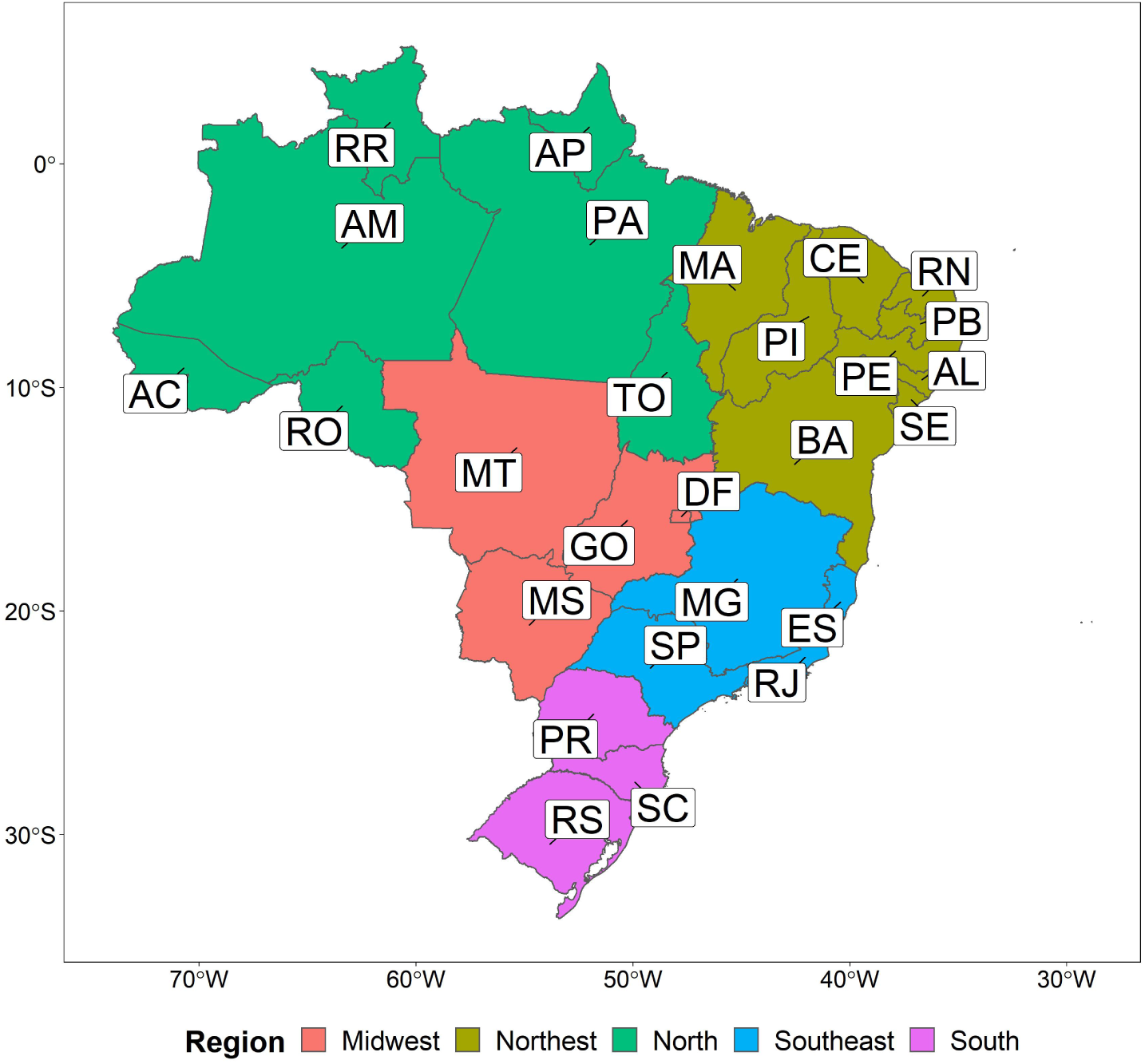
Map of Brazilian regions and states abbreviations.

The first case of COVID-19 in Latin America was confirmed on February 25 in a traveller returning to São Paulo city from the Lombardy region in Italy, the epicenter of the first wave of the disease in Europe, and the first death by the disease was reported on March 12, also in São Paulo. After that, with the epidemic evolving in Brazil, on March 20 the Ministry of Health recognized that community transmission was occurring across the country, as a strategy to ensure a collective effort and logistic and financial support for states and its population. This was followed by the implementation of nationwide non-pharmaceutical measures, including physical distancing, isolation, quarantine, compulsory notification, and mask-use mainly by elderly and exposed/infected people^10^.

Due to a lack of national coordination, the pandemic was mainly fought by the 27 states and 5,568 municipalities who took diverse, heterogeneous and mainly non-coordinated decisions by government officials at the federal, state and municipal levels. This led to both success and failure stories and to distinct efficacy of non-pharmaceutical measures in slowing the rate of transmission across the country, which outlined the importance of taking into account the socio-economical background of a region when implementing such measures. Such a diverse response to the pandemic is this paper reason of being, since in a such heterogeneous scenario a much needed assessment of the efficacy of the measures to stop the disease spread is not a straightforward task, so a careful quantitative and qualitative analysis of the disease evolution and of measures such as social isolation is needed to evaluate this efficacy.

This paper aims to asses the effectiveness of human mobility control measures on reducing the COVID-19 spread in Brazil, taking into account many aspects of the locations and their population, such as urban hierarchy, demography, and socio-economic profile. Our assessment focuses on the dataset containing daily cases of Severe Acute Respiratory Illness, and a daily isolation index, of the most important cities in Brazil in a period ranging from March 15 to October 30, 2020. We carefully dissect this dataset, coupling its quantitative analysis with qualitative information about measures enforced to slow the disease spread and characteristics of the population and locations around the country. With these analyses, we hope to give a snapshot of how the pandemic evolved in Brazil, and study the interplay between social isolation and COVID-19 spread in the country. Here we discuss and quantify in detail the conditions under which social distancing, measured through population local mobility, impact infection rates depending on different demography and socio-economic profiles. As a result, besides providing on its own insightful knowledge of the behavior of the pandemic in different conditions, our results allow models to be more precisely adjusted to take into account local characteristics and provide more reliable epidemic scenarios.

In Section 2 we present the material and methods of the quantitative analysis, while in Section 3 we present its results, and in Section 4 we discuss our findings.

## 2 Material and Methods

### 2.1 Data

#### 2.1.1 Human mobility

The social isolation index is a relative measure of the amount of people that do not leave their houses during the day. It can be calculated based on mobile users’ mobility data from many private companies that work with geolocation services^2,19,34,25^. The government of São Paulo state, in cooperation with the 4 main mobile network services in Brazil (Oi, Tim, Vivo, Claro), publicly provide an official social isolation index for more than 100 cities of the state. The mobile network companies use radio based geolocation methods to infer the position of users, which limits its applicability, being adequate only for large cities with many stations. Google^19^ and Apple^2^ only provide mobility data for a small subset of Brazilian cities.

Due to the low coverage of brazilian cities of Google, Apple and Network companies, in this study, we adopt a dataset provided by the company InLoco^∗25^. This company provides anonymous geolocation technologies for several mobile applications. The Software Development kit provided by InLoco uses multiple mobile sensors, including Wi-Fi, GPS, bluetooth and other, to infer the mobile location with the accuracy of meters. The company does not collect any user personal information and users need to *opt in* for geolocation services in the application. Several safety measures are used to ensure data safety and the company complies with Brazilian law of data protection, which ensures ethical and legal assurances of data collection and usage^26^.

For the social isolation index, the home location of an user is estimated based on the recent location registered during night periods. Once the home location is known for each user in the database, InLoco calculates the number of users that left their homes during the day. The house location and the breach in isolation is calculated considering an Hexagonal hierarchical geospatial indexing system (H3 - https://h3geo.org/) with resolution level 8 (hexagons have edges with approximately 460m of length). The H3 data is further aggregated to city level. For this work, only aggregated city level with the isolation index pre-computed from the company was available, therefore avoiding any anonymity issues in what concerns this work.

The main advantage of using the InLoco dataset is its wide coverage. In mid 2020 its database consisted of more than one fourth of the mobiles in the country ^†^. While this data can have sample biases, with respect to the nature of the mobile application use and smartphone diffusion in the country, its is, to our knowledge, the vastest dataset of this kind for Brazil. Also, it has been widely used in the pandemic to monitor public efforts to reduce mobility and also in several academic studies^5,6,14,32,33^. All data used in this work is provided as supplementary material.

In the analysis, for each municipality, the daily observed social isolation index was subtracted from the average value observed in the respective city from February 1 to February 15 (before carnival holiday and the start of the pandemic in Brazil). Therefore, in what follows, we will refer to the isolation index as being this difference from the reference period, interpreted as the *increment/reduction* in social isolation.

#### 2.1.2 Reported cases

The number of reported cases of Severe Acute Respiratory Illness (SARI), over time, was collected from the national database SIVEP-Gripe^11^. It comprises all severe hospitalized cases related to respiratory viruses whose notification is compulsory in Brazil. It has been a good thermometer to catch the disease spatio-temporal dynamics in the country, since it has struggled with an insufficient capacity of molecular diagnosis and fast tests^21^. The information available does not distinguish between imported and autochthonous cases. Although the first COVID-19 case occurred on February 25, the dataset used here ranges from March 15 until October 30, 2020, since before that changes were not yet implemented on the surveillance system of SIVEP-Gripe to identify COVID-19 cases among cases of other diseases such as Influenza, Respiratory Syncytial virus, Adenovirus, and Parainfluenza^4^. This period comprises the first wave of the disease in Brazil during which it is supposed the transmission of a unique variant of the virus in Brazilian territory.

A nowcasting procedure was performed, using the R package NobBS^30^, to correct delay in notifications which in Brazil can take up to forty days. Since the nowcasted daily number of cases still presented weekly variations, it was smoothed by taking a 7-day moving average. To compare the disease spread among the cities, the daily number of cases was divided by 100,000 inhabitants.

The effective reproduction number *R*_*t*_ was calculated using the nowcasted smoothed data of incidence. For this, we considered the epidemiological model SEIR (susceptible-exposed-infected-recovered), and the approach proposed by Wallinga and Lipsitch^36^. The parameters considered to calculate *R*_*t*_ are the latent period (*η*^−1^) of 3.0 days, the infectious period (*τ*^−1^) of 6.4 days, and the life expectancy in Brazil (*µ*^−1^) of 75 years. The rates of leaving the exposed and infectious classes are denoted by *s*_1_ = *η* + *µ* and *s*_2_ = *τ* + *µ*, respectively. Therefore, the generation interval distribution *g*(*t*) is given by^1^

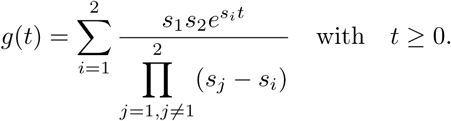

After normalizing *g*(*t*) we can evaluate *R*_*t*_ as

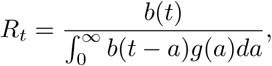

and the daily transmission index is calculated as *R*_*t*_ *× τ*.

#### 2.1.3 Urban hierarchy, human development index, and transportation infrastructure

One variable that can directly affect the spread of the epidemic over the country is the hierarchy of urban centers. In Brazil, the classification is done by Instituto Brasileiro de Geografia e Estatística (IBGE)^24^, and is based on territory management, offering of trade and services, financial services, offering of university education, media and communication markets, culture and sports, transportation services, agricultural activities, and international links. The urban hierarchy of Brazil has five levels: (i) Metropolises, (ii) Regional Capitals, (iii) Sub-Regional Centers, (iv) Zone Centers, and (v) Local Centers. The unit of this classification is the population arrangement consisting of the clustering of two or more municipalities and constitutes a reference framework of urbanization in the country. On each level, the main city that comprises the urban arrangement gives it its name.

The Metropolises level comprise the main urban centers of Brazil divided into three sub-levels: Greater National Metropolis, National Metropolis and Metropolis. The only Greater National Metropolis is São Paulo (SP), listed as an alpha global city by the Globalization and World Cities Research Network, with 21.5 million of inhabitants. The National Metropolises are the Federal District of Brasília (DF), with 3.9 million inhabitants, and Rio de Janeiro (RJ), which has a population of 12.7 million. These three cities are also the main destination of travel within and from outside of Brazil. Finally, the Metropolis sub-level is composed by: Belo Horizonte (MG), Curitiba (PR), Florianópolis (SC), Fortaleza (CE), Goiânia (GO), Manaus (AM), Belém (PA), Porto Alegre (RS), Recife (PE), Salvador (BA), Vitória (ES) and Campinas (SP). All of them but Campinas are capital cities of their respective states. The average population of Metropolises is 3 millions, with Belo Horizonte being the most populous with 5.2 million inhabitants and Florianópolis and Vitória the less populous with 1 and 1.8 million, respectively.

The second level, the Regional Capitals, are composed of 97 urban centers with regional influence, which are divided in three groups: A, B and C. The Regional Capitals A is composed of 8 capital cities of states in the Northeast and Midwest region, and the city of Ribeirão Preto (SP). These cities have a similar population size, varying between 0.8 and 1.4 million inhabitants, and share a direct relationship with Metropolises. There are 24 Regional Capitals B, ten of them in the South region, which have an average population of 530 thousand inhabitants, the most populous being São José dos Campos (SP) with 1.6 million inhabitants. These cities are reference centers within their states, with the exception of state capitals Palmas (TO) and Porto Velho (RO) which also exerts influence on other states. The Regional Capitals C are formed by 64 cities, 30 of them in the Southeast region, with average population size of 360 thousand inhabitants.

The Sub-regional Centers level is composed of 352 cities with less complex management activities than Regional Capitals. The forth level, Zone Centers, is composed of 398 cities with less management activities and, finally, the fifth level, the Local Centers, is composed of 4.037 cities, that is, 82.4% of urban centers of Brazil with an average of 12.5 thousand inhabitants and whose influence is restricted to its own territorial limits.

Socio-economic variables are also usually associated with disease spreading, and the Human Development Index (HDI) is known to be an important factor. The HDI combines measurements of life expectancy, education, and per-capita income. It ranges from 0,418 to 0,862 among the cities, while the states average ranges from 0,631 to 0,824. The index is generally greater on states in the center-south regions and lowest in the northern regions. Indeed, the higher average index can be found at São Paulo (SP), Santa Catarina (SC), and Distrito Federal (DF) and the lower at Pará (PA), Alagoas (AL), Maranhão (MA), and Piauí (PI).

Lastly, transportation infrastructure vary in the country. Although the primary source of transportation is the road transportation, it is highly concentrated in the center-south regions, specially in the state of São Paulo. The exception to this rule is the Amazon region where road transportation loses its importance to waterways thanks to the dense natural river network. The lack of adequate roads in the North region and the logistical issues with connecting roads with waterway transportation are some of the bottlenecks of the region’s development. From a human mobility point of view, while the majority of trips within states are by road, the main mean of transportation between states are airplanes, that count with a network of around 100 airports around the country. International airports are located at São Paulo (SP), Rio de Janeiro (RJ), Brasíla (DF), Belo Horizonte (MG), Campinas (SP), Salvador (BA), Fortaleza (CE), Recife (PE), Porto Alegre (RS), Florianópolis (SC), Manaus (AM), Belém (PA), Natal (RN), and Campo Grande (MS).

## 3 Results

### 3.1 Human mobility versus reported cases

In this section, we study the relationship between the daily social isolation index and the daily incidence of the disease in the main cities that comprise the metropolises, which are São Paulo, Brasília, Rio de Janeiro, Belo Horizonte, Curitiba, Florianópolis, Fortaleza, Goiânia, Manaus, Belém, Porto Alegre, Recife, Salvador, Vitória, and Campinas. Similar to what was done for the daily incidence, the daily social isolation index was smoothed by taking a 7-day moving average. Figure 2 shows the daily ratio between the social isolation index, and the daily incidence, and their respective daily average among the cities considered. Spatial-temporal variations of both measures highlight regional and temporal differences resulting from geographic, demographic, cultural and political characteristics of each main city of Brazil, whose behaviour affects other cities in their region of influence. In general, the social isolation index was negatively related with the incidence of the disease, but not linearly proportional. Besides, taking all cities together, the monthly average of the social isolation index decreased with time: from March to October they were (average value and its standard deviation), respectively, 0.21±0.02, 0.22±0.02, 0.18±0.03, 0.13±0.01, 0.13±0.02, 0.11±0.01, 0.09±0.01, and 0.08±0.01.

**Fig. 2.**
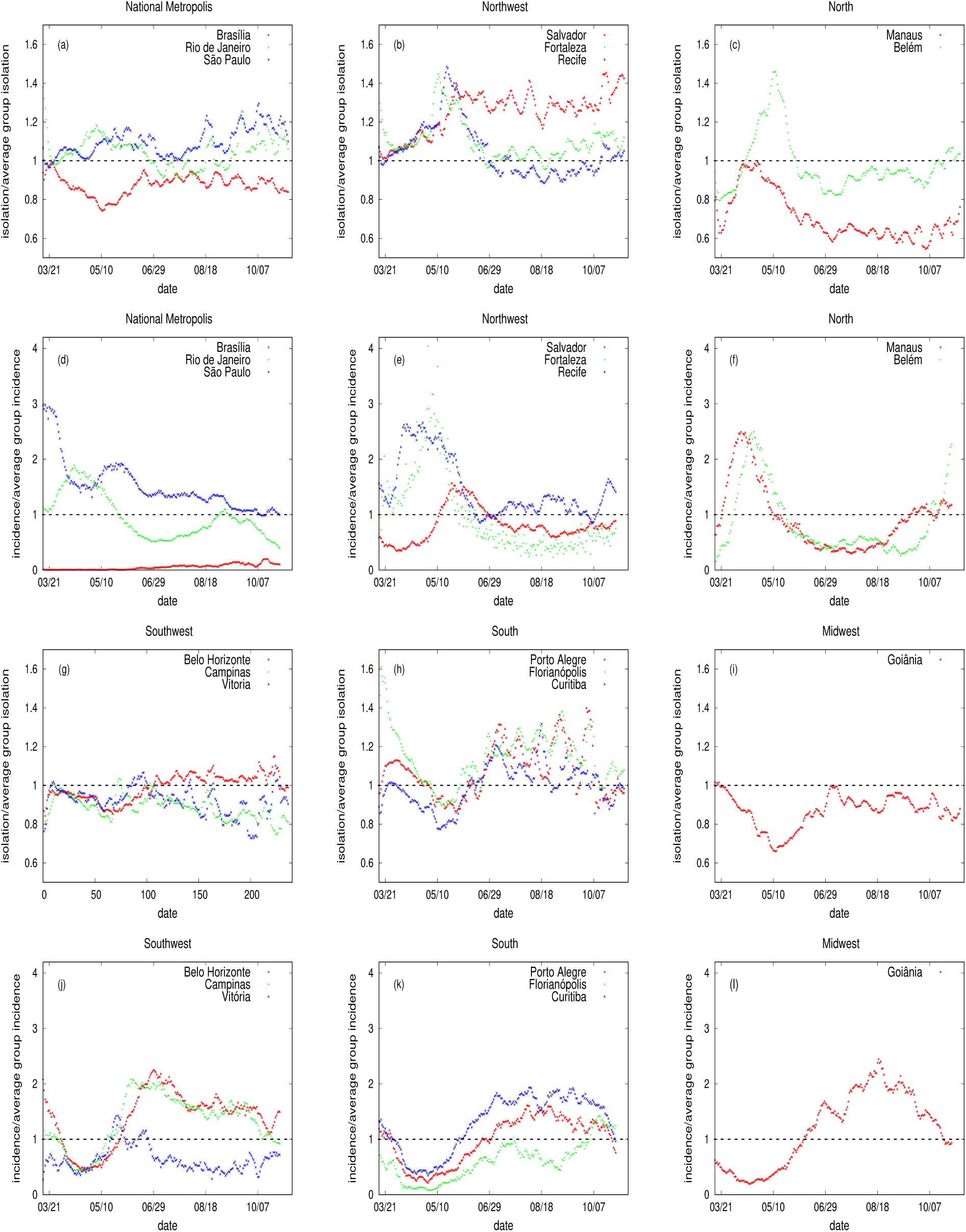
In (a-c), (g-i) the daily ratio between the social isolation index and its respectively daily average. In (d-f) and (j-l) the daily incidence and its respectively daily average. The average was calculated considering the main cities that represent each Metropolis. The date corresponds to the day of the first symptoms. The dashed horizontal line shows when this ratio is equal to 1.

In order to mitigate the effects of the disease on the healthcare system, lockdown strategies were implemented on some cities in the North and Northeast regions, causing the sharp increase on the social isolation index measured in Belém, Fortaleza and Recife (see Figure 2 around May 10). In Belém, differently from the other two cities, the lockdown can be clearly seen in the data since its social isolation index was below average in the period leading up to the lockdown and then increased way over the average. For these cities, the epidemic peak was from 2.5 to 4.2 times the observed average incidence on their respective days.

Among the cities of the Northeast region, there is Salvador, which seems to have kept a more efficient control of the disease transmission, since it endured a higher isolation rate, and had a smaller peak, compared to other cities of this region. On the other hand, there is Fortaleza, which has the lowest HDI among the three Northeast cities considered, and is one of the main entrance of travelers from outside the country visiting the North and Northeast regions, factors which can explain its low performance on controlling the spread of the disease relatively to the other two cities. Likewise, in the North region, despite the fact that the social isolation index was higher at Belém compared to Manaus, both cities had a similar epidemic temporal evolution pattern. Among the cities, these two cities have the lowest HDI and are highly connected by waterways. They belong to the Amazon region where the spread of COVID-19 took the course of the waterways and was enhanced by the long duration of the trips in boats, where mitigation strategies, such as socially distancing and hand washing, are compromised^22^.

In the Southeast region, although an increase on the social isolation index was observed in Belo Horizonte at the end of the study period, it had a similar transmission pattern as Campinas, while Vitória, which is among the cities with the highest HDI and lacks an international airport, had the best performance on disease control transmission when compared to the other Southeast cities, what can be due to these important factors. Among the South cities, despite Curitiba not having an international airport, its has worse performance on the social isolation index when compared to Porto Alegre and Florianópolis, which may be behind its larger number of reported cases. Still in the South, it is worth mentioning that not only Florianópolis has a high HDI, but also a major part of its territory is an island, which could have contributed to its high performance on controlling the disease spreading when compared to the other South cities.

When comparing both cities of the Midwest region, one sees that Goiânia had a social isolation index pattern similar to Brasília, but its performance to contain the spread of the disease was worse. The fact that Brasília has a higher HDI and lower population density may explain the low disease incidence compared to the other cities. Another important factor is that the Midwest region annually records the lowest number of influenza and other respiratory viruses cases inside the national territory^13^, so these cities might be prone to have lower incidence of a respiratory disease. Lastly, Rio de Janeiro and São Paulo, both Southeast cities, are highly connected by roadways and airways, and share a similar pattern of social isolation index, with São Paulo, in general, having a better performance. Differently from the others cities, São Paulo has been having an outbreak of COVID-19 always above the average.

### 3.2 Lockdown strategy

Strict lockdown strategies were not common in Brazil in 2020, but were nonetheless implemented in four capital cities of states in the northern regions. In Figure 3 we present the daily incidence (in brown) and the *R*_*t*_ (in blue) for four cities which implemented a lockdown, namely, São Luís, Belém, Fortaleza and Recife. Among the four cities, the first confirmed case occurred in Recife on March 12, which was then followed by Fortaleza on March 15, Belém on March 18, and São Luís on March 20. The disease spread differently in each city, which also have distinct hospital and test capacities. The first city to declare lockdown was São Luís on May 5 when it achieved 10.75 new cases per day (average of the 7 days before lockdown). It was then followed by Belém on May 7 with 7.14 new cases per day, Fortaleza on May 8 with 11.35 new cases per day, and Recife on May 16 with 8.17 new cases per day. In São Luís, the lockdown took 13 days, and it was observed a reduction of 32.5% on the number of cases per day. In Belém it took 18 days with a reduction of 19.8% on the number of cases per day. In Fortaleza it took 24 days with a reduction of 33.6% of the cases per day. Finally, in Recife it took 16 days with a reduction of 8% on the number of cases per day.

**Fig. 3.**
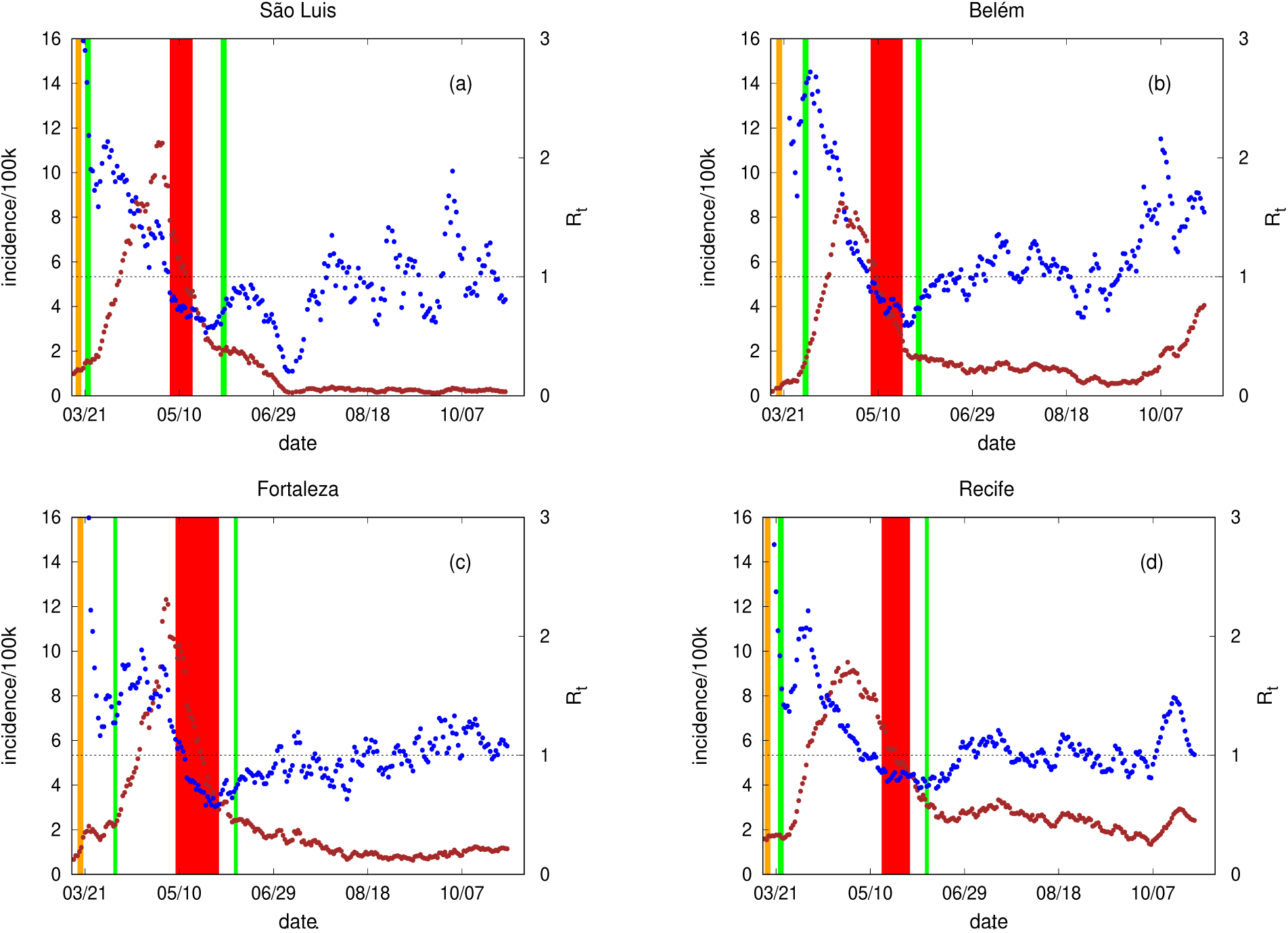
Temporal evolution of incidence and *R*_*t*_, respectively in brown and blue. In orange, red and green, we have the start of control measurements, the lockdown and some degree of flexibilization, respectively. In (a) we have São Luís, (b) Belém, (c) Fortaleza, and (b) Recife. The date corresponds to the day of the first symptoms. The dashed horizontal line is drawn for *R*_*t*_ = 1.

In Figure 3, the orange vertical band represent the declaration of an emergency situation, characterized by the closing of schools, universities, bars, shopping malls, commerce, etc. In green we had the flexibilization of some measurements, and in red the lockdown period. Table 1 shows different measures on the data calculated before, during and after the lockdown period. This analysis was done considering a lag of seven days (*≈* incubation period) between the social isolation index and the daily incidence.

**Table 1.** Percentage variation of the social isolation index and the transmission index per period. From the first to the fourth, period means week. In the case of Fortaleza and Recife, the fifth period is also a week. For all of them, the second week corresponds to the lockdown. In all cases, the last period is counted until the first minimum of the incidence curve after the lockdown period is finished. A lag of seven days between the isolation index and the transmission index was considered.

| São Luis |  | Belém |  | Fortaleza |  | Recife |  | Period |
| --- | --- | --- | --- | --- | --- | --- | --- | --- |
| isol. index | trans. index | isol. index | trans. index | isol. index | trans. index | isol. index | trans. index | - |
| -0.0031 | 0.0176 | -0.0164 | -0.1907 | -0.0196 | -0.1368 | 0.0484 | -0.0758 | 1 |
| **0.1134** | **-0.3595** | **0.0318** | **-0.2165** | **0.1303** | **-0.2813** | **0.1529** | **-0.1278** | **2** |
| -0.0533 | -0.1035 | 0.0115 | -0.1293 | -0.0471 | -0.2425 | -0.1080 | 0.0300 | 3 |
| -0.2704 | -0.1086 | -0.1722 | -0.1130 | -0.0927 | -0.1462 | -0.1995 | -0.0410 | 4 |
| -0.1716 | -0.0513 | -0.1800 | -0.0945 | -0.1632 | 0.0986 | -0.1562 | -0.0758 | 5 |
| - | - | - | - | -0.1544 | 0.0574 | -0.0545 | -0.0058 | 6 |

In all cases, the lockdown was effective on reducing the incidence (and *R*_*t*_ values). This control method was clearly efficient in São Luís, which was presenting an increase on the rate of transmission before lockdown that changed to a decreasing pattern after it. Fortaleza, Belém and Recife were already diminishing the rate of transmission before lockdown occurred. Among them, Fortaleza was able to keep it longer with a high social isolation index. Although having a huge increase on social isolation index as a result of lockdown, Recife was not able to keep it for long. As a consequence, the *R*_*t*_ value after lockdown was similar to the one measured before it. It is interesting to note that in Recife the social isolation index was already increasing before lockdown.

For São Luís, the maximum incidence occurred six days before lockdown, and after lockdown disease incidence still decreased until fifteen days after it ended. For Belém, the maximum incidence occurred sixteen days before lockdown, and after lockdown the incidence still decreased until five days after it ended. For Fortaleza, the maximum incidence occurred five days before lockdown, and after lockdown the incidence still decreased until ten days after it ended. For Recife, the maximum incidence occurred eighteen days before lockdown, and after lockdown the incidence still decreased until fifteen days after it ended. The highest percentage on the reduction of the infection observed in São Luís can be related to its lower position on the urban hierarchy when compared to the other three, and to its spatial geography (it is an island). All these cities have public laboratories for the molecular diagnosis of SARS-CoV-2, but we can not compare the diagnostic capacity of each one because this data is not publicly available. The lack of diagnostic capacity can jeopardize control efforts by causing delay on decision making.

### 3.3 São Paulo mitigation strategies

The history of SARS-Cov-2 spreading on the State of São Paulo is characterized by different time and spatial scales. Starting from the regional centers, the disease displaced to municipalities with major connections, after to municipalities with minor connections, and lastly to rural municipalities (spatial pattern well explained by the gravity model). Locally, the spread was by contiguity (diffusion model)^16^. As the disease spread, following the example of the metropolitan region of São Paulo, the inner state adopted strong mitigation measures closing schools, universities, and all its trade, keeping only essential services such as pharmacies, supermarkets, and hospitals open. This delayed the arrival of the virus to the inner state by at least one month, letting the cities prepare their healthcare systems, which are more fragile in this region.

On May 27, São Paulo State started to move out from a restrictive quarantine with a flag system that classifies, based on several indicators, the disease transmission risk, and the probability of break-down of the healthcare system. Five colors were adopted: (i) red (phase 1) is considered a contamination phase and only essential services are permitted; (ii) orange (phase 2) is considered an attention phase, with the possibility of some services opening (limit of 20% of its capacity and maintaining all specific hygiene protocols); (iii) yellow (phase 3) is considered a controlled phase, with some flexibilization (limit of 40% of its capacity and maintaining all specific hygiene protocols); (iv) green (phase 4) is a partial opening phase, in which all services are allowed to open (limit of 60% of its capacity and maintaining all specific hygiene protocols); (v) blue (phase 5) has only restrictions over events that generate large agglomeration of people. Restaurants, fitness centers, cultural activities, and beauty salons are allowed from phase 3. At phase 4, schools and universities can be opened with 35% of the classes occupied (Plano São Paulo, https://www.saopaulo.sp.gov.br/planosp/). These phases were attributed to each region of the State, defined according to a division of its healthcare system^12^. These regions, called Regional Health Departments (DRS, in portuguese), are each one represented by a major centralized city: Araraquara, Araçatuba, Baixada Santista, Barretos, Bauru, Campinas, Franca, São Paulo Metropolitan Area, Marília, Piracicaba, Presidente Prudente, Registro, Ribeirão Preto, São João da Boa Vista, São José do Rio Preto, Sorocaba, and Taubaté.

Figure 4 shows the temporal evolution of the incidence (in brown) and the *R*_*t*_ (in blue) for selected cities in São Paulo State. The relation between the transmission index and the social isolation index is also displayed (with a lag of seven days between them). The other colors (vertical bands) are associated with each moment of the São Paulo’s Plan in each city. The peak of the incidence curve occurred on May 5 at São Paulo (with 6 new cases) on June 10 at Campinas (with 5.9 new cases), and on August 10 at Votuporanga (with 8.5 new cases). This pattern shows the spread of the disease from the metropolis of São Paulo to inner cities in a lower degree of urban hierarchy. Besides, less complex cities (less level of urban hierarchy compared to São Paulo) were not able to achieve a high social isolation index or keep it for a long time. The variance observed on the transmission rate can be related to the population density in each city which is respectively 7398.3, 1359.6, and 201.2 inhab/km^2^ at São Paulo, Campinas, and Votuporanga. Although the incidence was higher at Votuporanga, the average absolute number of cases was 3.4 (from 1 to 11), 36.3 (from 9 to 95), and 422.8 (from 161 to 986) in Votuporanga, Campinas, and São Paulo, respectively.

**Fig. 4.**
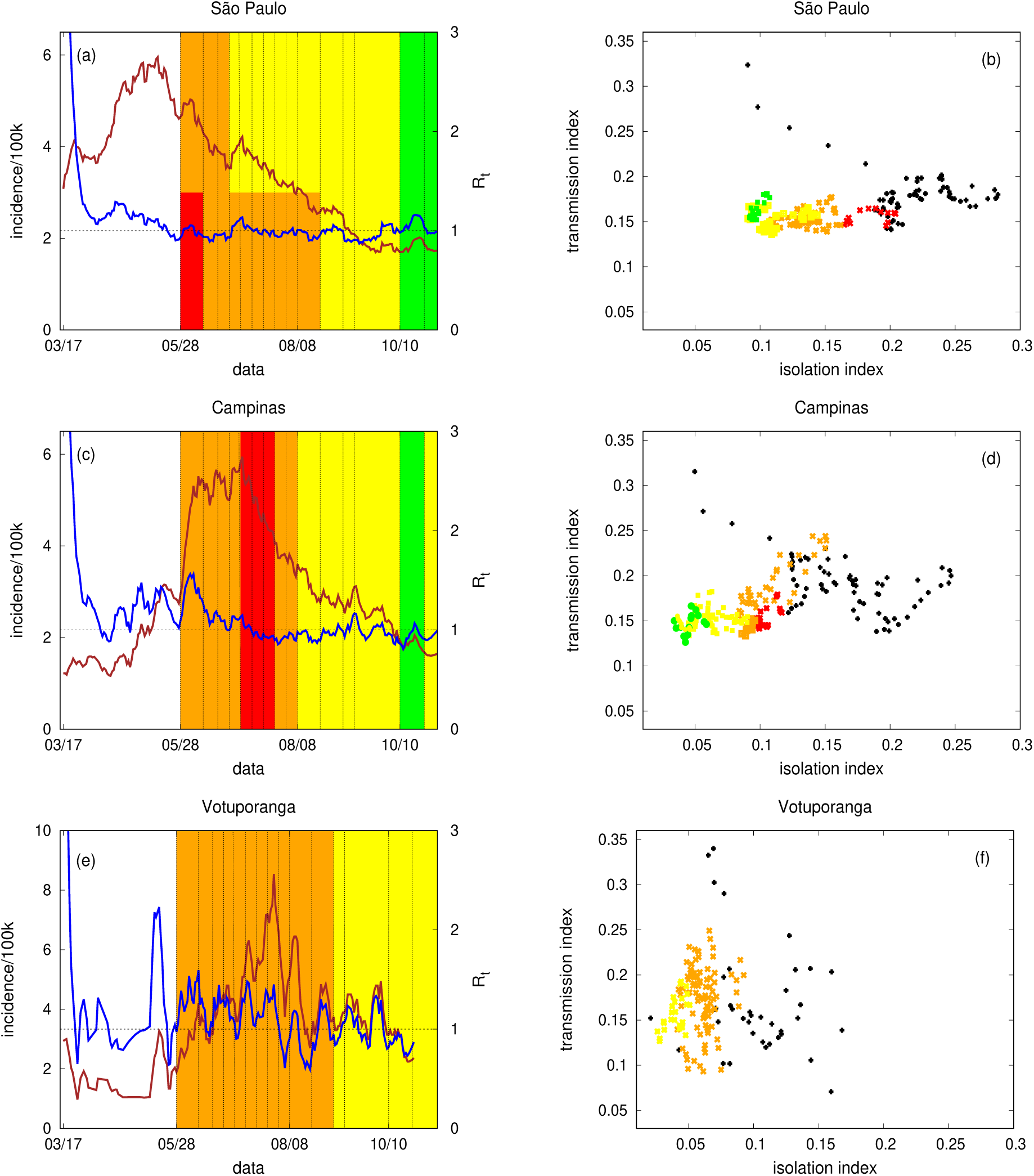
São Paulo (DRS I), Campinas (DRS XII), and Votuporanga (DRS XV). In (a), (c), (e) the temporal evolution of incidence and *R*_*t*_, respectively in brown and blue. In (b), (d), (f) the isolation index versus the transmission index (with a lag of seven days between them). The colors in all panels represent the levels of São Paulo plan. The date corresponds to the day of the first symptoms. The (daily social) isolation index was subtracted from the average value observed from February 1 to February 15, and the (daily) transmission index is calculated as *R*_*t*_ *× τ* where *τ*^−1^ is the infectious period.

### 3.4 Relation between social isolation index and the speed of the disease dispersion

In this section, we study the relationship between social isolation index and how fast the disease spread over different geographic locations around the country. The enrolled cities satisfy one of the following criteria: (i) is the capital of a state, (ii) is the first or second city of a state with a higher number of cases, (iii) belongs to São Paulo state and is among its cities with highest number of cases.

For each municipality, the smoothed curve of daily cases in the upward phase, that is, from March 15 until the day when the maximum number of cases occurred, was considered. Then, the upward phase was divided into seven stages in the following way. First, we calculated the total number of infected people in this period (upward phase), divided it in half and marked the day in which such a number of cases was reached as the start of stage 7, which ends when the peak of cases is reached. Therefore, stage 7 represents the period contemplating the second half of the cases until the peak. Analogously, new successive divisions were made, minding the day when a corresponding half of accumulated cases was reached. In total, six divisions were made, creating 7 stages. In this way, stage 1 refers to period between March 15 and the occurrence of 1.5625% of the cases in the upward phase; stage 2 refers to period between 1.5625% and 3.125% of it; and stages 3 to 6 correspond to periods between 3.125%-6.25%, 6.25%-12.5%, 12.5%-25%, 25%-50%, respectively, of cases in the upward phase (see Figure 5).

**Fig. 5.**
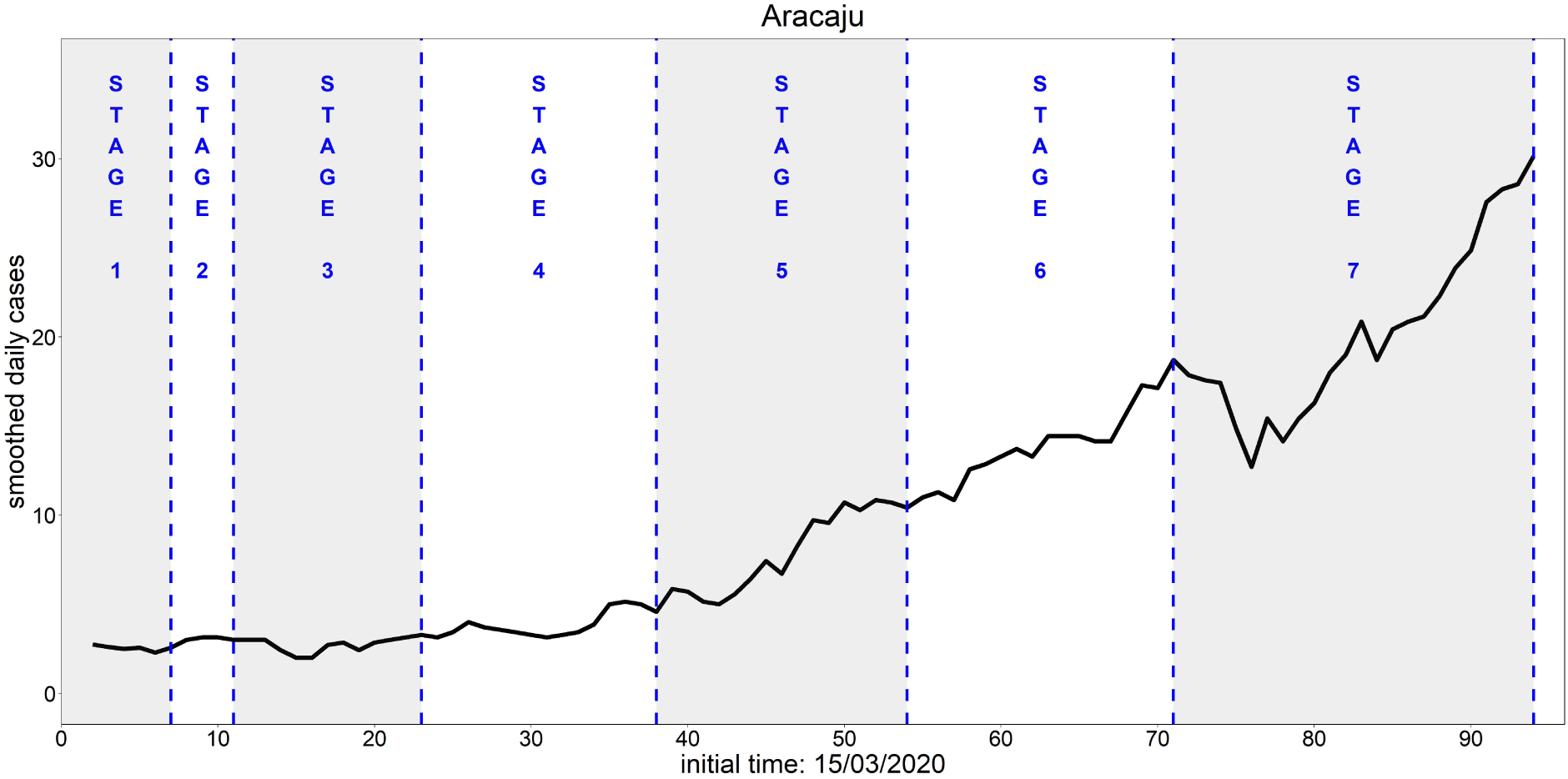
Temporal evolution of the number of cases for the municipality of Aracaju. The date corresponds to the day of the first symptoms, measured from (t=0) March, 15. The stage 1 refers to period between March 15 and the occurrence of 1.5625 % of the cases in the upward phase and the stage 2 refers to period between 1.5625% and 3.125% of it. The stages 3 to 7 correspond to periods between 3.125%-6.25%, 6.25%-12.5%, 12.5%-25%, 25%-50% and 50%-100% of cases in the upward phase, respectively.

The length of these stages is an estimate of the speed of disease spreading. When efforts to stop or diminish the disease spread are implemented in one stage, one expects an increase in the length of later stages. Hence, finding a relationship between the efforts to control the disease spread, such as the social isolation index, and the length of these stages could be an efficient way of assessing the efficacy of the control measures.

In Table 2 we present, for each municipality (a total of 31 analyzed), the median of the social isolation index and the length of the disease spreading stages. The incidence was associated to the social isolation index considering a lag of seven days. The social isolation index at day *t*− 7 is the average between the ones observed at *t* − 8, *t* − 7 and *t* − 6. In bold, are highlighted the values of the social isolation index above the third quartile (Q3) and the length of the stages (*Δ*_*i*_ with *i* = 1, 2, …, 7) which are above the respective median.

**Table 2.**
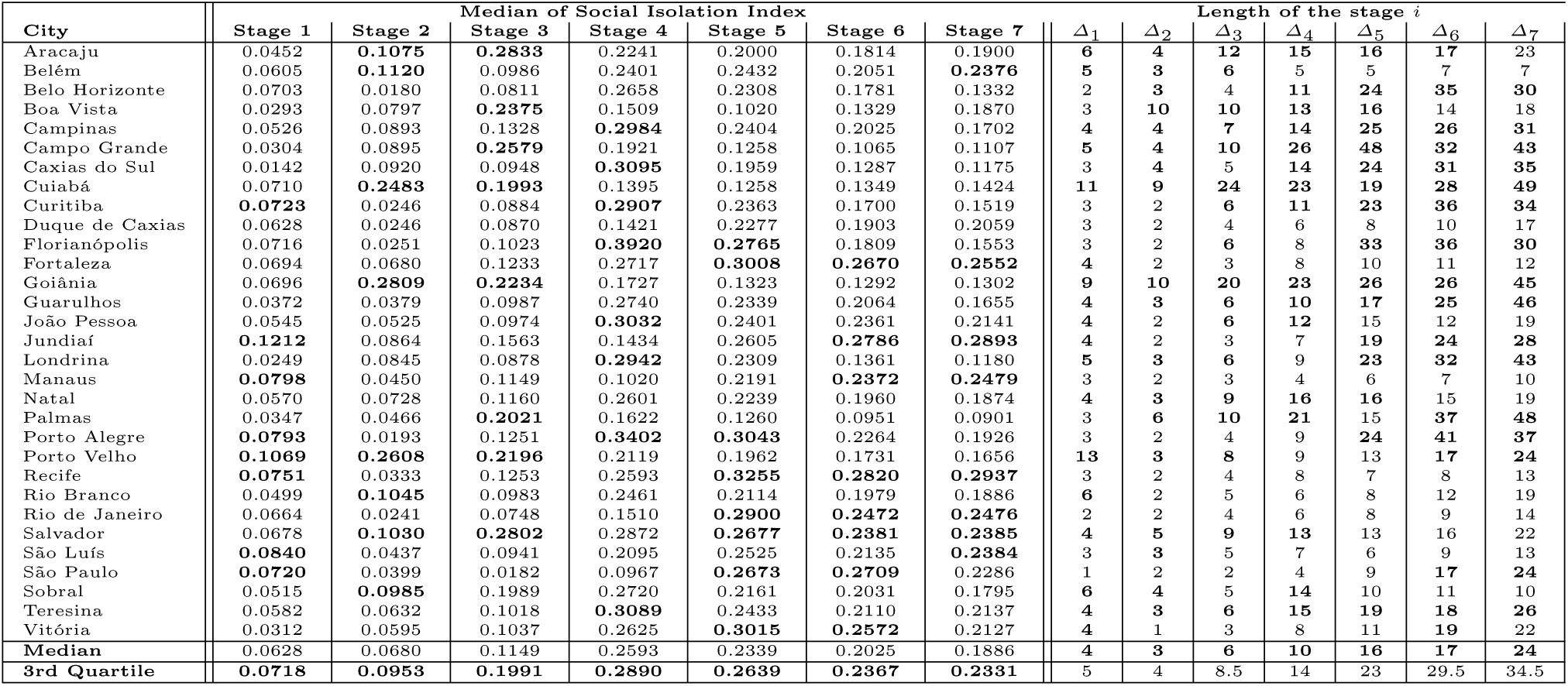
Median of social isolation index and length of the stages (*Δ*_*i*_ with *i* = 1, 2, 3, 4, 5, 6, 7) of the upward phase for each municipality. The stage 1 refers to period between March 15 and the occurrence of 1.5625 % of the cases in the upward phase and the stage 2 refers to period between 1.5625% and 3.125% of it. The stages 3 to 7 correspond to periods between 3.125%-6.25%, 6.25%-12.5%, 12.5%-25%, 25%-50% and 50%-100% of cases in the upward phase, respectively.

| City | Median of Social Isolation Index | | | | | | | Length of the stage $i$ | | | | | | |
| --- | --- | --- | --- | --- | --- | --- | --- | --- | --- | --- | --- | --- | --- | --- |
| | Stage 1 | Stage 2 | Stage 3 | Stage 4 | Stage 5 | Stage 6 | Stage 7 | $\Delta_1$ | $\Delta_2$ | $\Delta_3$ | $\Delta_4$ | $\Delta_5$ | $\Delta_6$ | $\Delta_7$ |
| Aracaju | 0.0452 | **0.1075** | **0.2833** | 0.2241 | 0.2000 | 0.1814 | 0.1900 | **6** | **4** | **12** | **15** | **16** | **17** | 23 |
| Belém | 0.0605 | **0.1120** | 0.0986 | 0.2401 | 0.2432 | 0.2051 | **0.2376** | **5** | **3** | **6** | 5 | 5 | 7 | 7 |
| Belo Horizonte | 0.0703 | 0.0180 | 0.0811 | 0.2658 | 0.2308 | 0.1781 | 0.1332 | 2 | **3** | 4 | **11** | **24** | **35** | **30** |
| Boa Vista | 0.0293 | 0.0797 | **0.2375** | 0.1509 | 0.1020 | 0.1329 | 0.1870 | 3 | **10** | **10** | **13** | **16** | 14 | 18 |
| Campinas | 0.0526 | 0.0893 | 0.1328 | **0.2984** | 0.2404 | 0.2025 | 0.1702 | **4** | **4** | **7** | **14** | **25** | **26** | **31** |
| Campo Grande | 0.0304 | 0.0895 | **0.2579** | 0.1921 | 0.1258 | 0.1065 | 0.1107 | **5** | **4** | **10** | **26** | **48** | **32** | **43** |
| Caxias do Sul | 0.0142 | 0.0920 | 0.0948 | **0.3095** | 0.1959 | 0.1287 | 0.1175 | 3 | **4** | 5 | **14** | **24** | **31** | **35** |
| Cuiabá | 0.0710 | **0.2483** | **0.1993** | 0.1395 | 0.1258 | 0.1349 | 0.1424 | **11** | **9** | **24** | **23** | **19** | **28** | **49** |
| Curitiba | **0.0723** | 0.0246 | 0.0884 | **0.2907** | 0.2363 | 0.1700 | 0.1519 | 3 | 2 | **6** | **11** | **23** | **36** | **34** |
| Duque de Caxias | 0.0628 | 0.0246 | 0.0870 | 0.1421 | 0.2277 | 0.1903 | 0.2059 | 3 | 2 | 4 | 6 | 8 | 10 | 17 |
| Florianópolis | 0.0716 | 0.0251 | 0.1023 | **0.3920** | **0.2765** | 0.1809 | 0.1553 | 3 | 2 | **6** | 8 | **33** | **36** | **30** |
| Fortaleza | 0.0694 | 0.0680 | 0.1233 | 0.2717 | **0.3008** | **0.2670** | **0.2552** | **4** | 2 | 3 | 8 | 10 | 11 | 12 |
| Goiânia | 0.0696 | **0.2809** | **0.2234** | 0.1727 | 0.1323 | 0.1292 | 0.1302 | **9** | **10** | **20** | **23** | **26** | **26** | **45** |
| Guarulhos | 0.0372 | 0.0379 | 0.0987 | 0.2740 | 0.2339 | 0.2064 | 0.1655 | **4** | **3** | **6** | **10** | **17** | **25** | **46** |
| João Pessoa | 0.0545 | 0.0525 | 0.0974 | **0.3032** | 0.2401 | 0.2361 | 0.2141 | 4 | 2 | **6** | **12** | 15 | 12 | 19 |
| Jundiaí | **0.1212** | 0.0864 | 0.1563 | 0.1434 | 0.2605 | **0.2786** | **0.2893** | **4** | 2 | 3 | 7 | **19** | **24** | **28** |
| Londrina | 0.0249 | 0.0845 | 0.0878 | **0.2942** | 0.2309 | 0.1361 | 0.1180 | **5** | **3** | **6** | 9 | **23** | **32** | **43** |
| Manaus | **0.0798** | 0.0450 | 0.1149 | 0.1020 | 0.2191 | **0.2372** | **0.2479** | 3 | 2 | 3 | 4 | 6 | 7 | 10 |
| Natal | 0.0570 | 0.0728 | 0.1160 | 0.2601 | 0.2239 | 0.1960 | 0.1874 | **4** | **3** | **9** | **16** | **16** | 15 | 19 |
| Palmas | 0.0347 | 0.0466 | **0.2021** | 0.1622 | 0.1260 | 0.0951 | 0.0901 | 3 | **6** | **10** | **21** | 15 | **37** | **48** |
| Porto Alegre | **0.0793** | 0.0193 | 0.1251 | **0.3402** | **0.3043** | 0.2264 | 0.1926 | 3 | 2 | 4 | 9 | **24** | **41** | **37** |
| Porto Velho | **0.1069** | **0.2608** | **0.2196** | 0.2119 | 0.1962 | 0.1731 | 0.1656 | **13** | **3** | **8** | 9 | 13 | **17** | **24** |
| Recife | **0.0751** | 0.0333 | 0.1253 | 0.2593 | **0.3255** | **0.2820** | **0.2937** | 3 | 2 | 4 | 8 | 7 | 8 | 13 |
| Rio Branco | 0.0499 | **0.1045** | 0.0983 | 0.2461 | 0.2114 | 0.1979 | 0.1886 | **6** | 2 | 5 | 6 | 8 | 12 | 19 |
| Rio de Janeiro | 0.0664 | 0.0241 | 0.0748 | 0.1510 | **0.2900** | **0.2472** | **0.2476** | 2 | 2 | 4 | 6 | 8 | 9 | 14 |
| Salvador | 0.0678 | **0.1030** | **0.2802** | 0.2872 | **0.2677** | **0.2381** | **0.2385** | **4** | **5** | **9** | **13** | 13 | 16 | 22 |
| São Luís | **0.0840** | 0.0437 | 0.0941 | 0.2095 | 0.2525 | 0.2135 | **0.2384** | 3 | **3** | 5 | 7 | 6 | 9 | 13 |
| São Paulo | **0.0720** | 0.0399 | 0.0182 | 0.0967 | **0.2673** | **0.2709** | 0.2286 | 1 | 2 | 2 | 4 | 9 | **17** | **24** |
| Sobral | 0.0515 | **0.0985** | 0.1989 | 0.2720 | 0.2161 | 0.2031 | 0.1795 | **6** | **4** | 5 | **14** | 10 | 11 | 10 |
| Teresina | 0.0582 | 0.0632 | 0.1018 | **0.3089** | 0.2433 | 0.2110 | 0.2137 | **4** | **3** | **6** | **15** | **19** | **18** | **26** |
| Vitória | 0.0312 | 0.0595 | 0.1037 | 0.2625 | **0.3015** | **0.2572** | 0.2127 | **4** | 1 | 3 | 8 | 11 | **19** | 22 |
| Median | 0.0628 | 0.0680 | 0.1149 | 0.2593 | 0.2339 | 0.2025 | 0.1886 | **4** | **3** | **6** | **10** | **16** | **17** | **24** |
| 3rd Quartile | **0.0718** | **0.0953** | **0.1991** | **0.2890** | **0.2639** | **0.2367** | **0.2331** | 5 | 4 | 8.5 | 14 | 23 | 29.5 | 34.5 |

Comparing the length of stages 5, 6, and 7 with their respective medians, we see that 24 municipalities (77.4%) maintain the same behavior, that is, they have the length of stages 5, 6, and 7 above the median or below the median in all three stages. Also, the correlation coefficients between the length of these stages are 0.773 between stages 5 and 6, 0.691 between stages 5 and 7, and 0.849 between stages 6 and 7. Due to the similarity of these stages, plus the fact that they correspond to 87.5% of all cases in the upward phase, our analysis will be restricted to stage 7.

Table 3 shows that a high social isolation index in stages 3 and 4, can slow the transmission of the disease in stage 7. From these 16 cities with length at stage 7 above the median, 12 (75%) of them had isolation above Q3 in stages 3 or 4. The four cities that did not had isolation above Q3 in these two stages were: Belo Horizonte, Guarulhos, Jundiaí and São Paulo. In the case of São Paulo, the length of the initial stages of disease spreading in this city was extremely short, indicating a fast initial dispersion of the disease. A social isolation index above Q3 was only observed in stages 5 and 6. This late high social isolation index may decrease the speed of disease, leading to the length of the final stages above of median. The city of Jundiaí had social isolation index above Q3 only in stages 1, 6 and 7, but presented a social isolation index above median in stages 2, 3 and 5. Therefore, except for stage 4, which was short (7 days), Jundiaí presented a social isolation index above the median in all the stages, which may have contributed to the reduction of the speed of disease transmission. On the other hand, we have the city of Guarulhos, that had a social isolation index above or equal the median in the stages 4, 5 and 6, and Belo Horizonte that had social isolation index above of median in stage 4 and really close to the median in stage 5, which may have contributed to a duration superior than the median in the stage 7.

**Table 3.** Distribution of municipalities according to social isolation index and disease transmission speed. The stage 1 refers to period between March 15 and the occurrence of 1.5625 % of the cases in the upward phase and the stage 2 refers to period between 1.5625% and 3.125% of it. The stages 3 to 7 correspond to periods between 3.125%-6.25%, 6.25%-12.5%, 12.5%-25%, 25%-50% and 50%-100% of cases in the upward phase, respectively. *Δ*_7_ is the lenght of stage 7 and *Q*3 is the third quartile.

|  | Social Isolation Index above Q3 |  |
| --- | --- | --- |
|  | Stages 3 or 4 | Stages 1, 2, 5, 6 or 7 |
| Municipalities with $\Delta_7$ below or equal to median | Aracaju<br>Boa Vista<br>João Pessoa<br>Salvador | Belém<br>Fortaleza<br>Manaus<br>Recife<br>Rio Branco<br>Rio de Janeiro<br>São Luís<br>Sobral<br>Vitória |
| Municipalities with $\Delta_7$ above or equal to median | Campinas<br>Campo Grande<br>Caxias do Sul<br>Cuiabá<br>Curitiba<br>Florianópolis<br>Goiânia<br>Londrina<br>Palmas<br>Porto Alegre<br>Porto Velho<br>Teresina | Jundiaí<br>São Paulo |

\*The municipalities of Belo Horizonte, Duque de Caxias, Guarulhos and Natal did not have an isolation index above the third quartile at any stage

Table 2 and 3 also present the cities at which the stage 7 length was below the median. From theses 15 cities, 11 (73.3%) did not have social isolation index above Q3 in stages 3 or 4. The four cities which presented social isolation index above Q3 were: Aracaju, Boa Vista, João Pessoa and Salvador. Note that João Pessoa, Aracaju and Salvador had length of stage 7 equal to 19, 23 and 22 days, respectively, values which are close to the median 24. The city Boa Vista had social isolation index above Q3 in stage 3, however, it had social isolation index below first quartile (Q1) in stages 4, 5 and 6, which has contributed for the short duration of 18 days obtained in stage 7.

The city that peaked faster was Manaus (35 days), followed by Belém (38 days), Recife and Rio de Janeiro (45 days), São Luiz (46 days), Duque de Caxias and Fortaleza (50 days). Among these cities, we can observe that Fortaleza kept the social isolation index above the median in all the stages. This bad performance to control the epidemics despite the high social isolation index can be related to the fact that this city is the main entrance of individuals coming from outside of the country to the North and Northwest of Brazil. Also the bad performance of Duque de Caxias can be explained by the fact that they are in the region of Rio de Janeiro influence.

### 3.5 Daily number of cases and the social isolation index during the upward phase

In this section, we evaluate, in detail, how the daily incidence is related to the social isolation index during the upward phase of the epidemic. Here, we divided the upward phase of the epidemiological curve into intervals. In each interval, we have either an increase or a decrease of the incidence. We defined an increase (or decrease) in the incidence if it is increasing (or decreasing) on the extreme points of the interval and we have at upmost two consecutive decreases (or increases) in the interior of the interval. The starting point, for each epidemic curve, corresponds to the calendar day at which 8.113 accumulated cases per 100k inhabitants were achieved. This threshold was chosen based on the epidemic growth observed in São Paulo city since this value is equivalent to 1000 accumulated cases in this city. In this analysis, we consider all cities not under the direct influence of São Paulo^†^, with *Δ*_7_ above or equal to the median, and social isolation index above Q3 in stages 3 or 4 (see Table 3), which were the cities with earlier high isolation, and a slower stage 7, which could be caused in part by this high isolation.

As an example, we show in Figure 6 the daily incidence for the city of Porto Alegre where the days in which the incidence increases or decreases are highlighted in different colors. Besides the temporal evolution of the incidence, we plotted the daily social isolation index against the daily incidence. Several features can be observed in this figure. A higher social isolation index was observed at the beginning of the epidemic, which seems to have promoted an efficient control of the incidence. With time, the isolation index decreased, which may have caused the epidemic curve to oscillate as can be seen by the large number of colors in the figure until the 70th day. Bellow some threshold of the social isolation index (around 0.16) a sharp vertical increase of the incidence can be seen, that is the days in blue. The increase of the social isolation index after this explosion of cases does not seem to control of the epidemic. This is an indicative that maybe if the social isolation index was kept 0.16 unit above of the reference value measured in February, Porto Alegre could have avoided the explosion of cases observed after the 70th day.

**Fig. 6.**
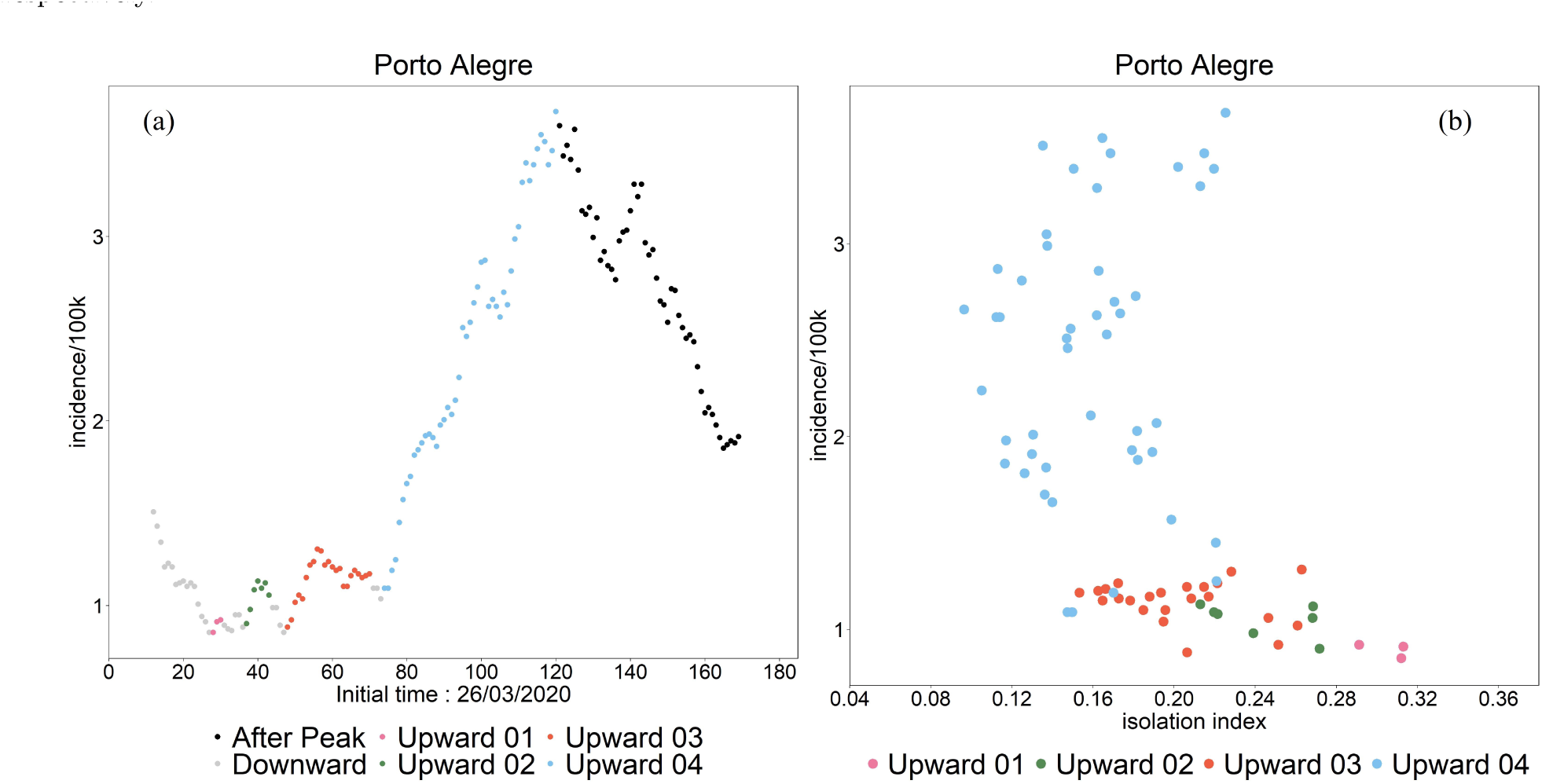
(a) Temporal evolution of the daily incidence and (b) the dispersion between the daily incidence and the isolation index for the city of Porto Alegre. The colors refer to periods of days where the incidence increases or decreases for at least three in a row. The downward phase of the incidence curve is in black.

The dispersion plot between the daily incidence and social isolation, colored according to the increase or decrease in cases, is presented in Figures 7 and 8. Five of those cities are Metropolises, three are Regional Capitals A and four are Regional Capitals B. A similar behavior was observed for the cities considered which have implemented some control measures in stages 3 or 4 (section 3.4) and are not under the influence of a Metropolis that did not do some mitigation control in these stages. For all these cities there seems to be a threshold, which may vary from city to city, where a decrease on the isolation index triggers an abrupt increase on the daily incidence. This phenomenon is characterized by the *L*-shape of the dispersion between the isolation index and daily incidence, where the horizontal part of the *L* refers to a period when higher isolation may have held the incidence low, and the vertical part of the *L* refers to a period when either the variation on the isolation does not affect the incidence or the decrease on the isolation below the threshold was followed by an explosion of cases.

**Fig. 7.**
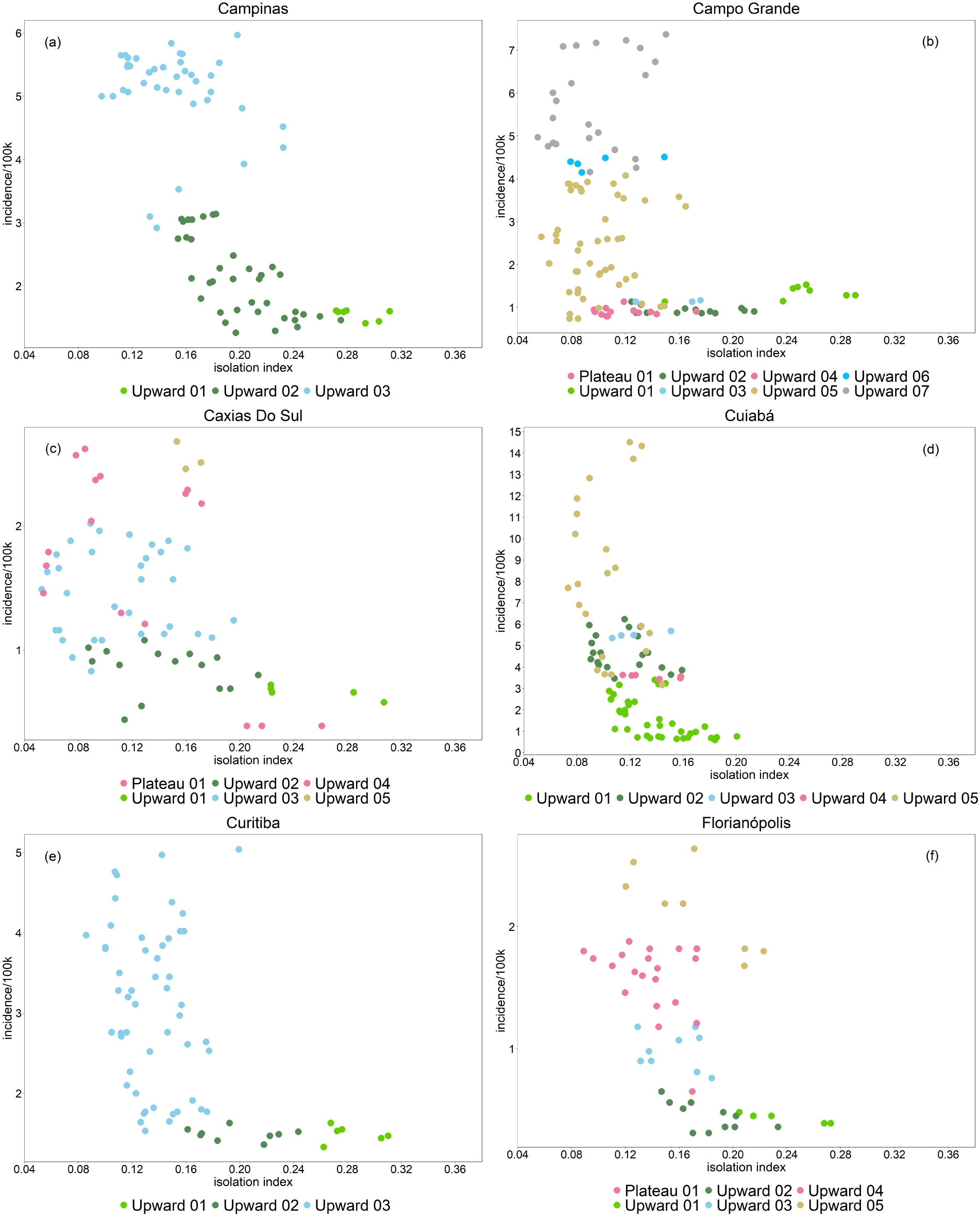
Dispersion between the daily incidence and the isolation index for the cities of (a) Campinas-SP, (b) Campo Grande-MS, (c) Caxias do Sul-RS, (d) Cuiabá-MT, (e) Curitiba-PR and (f) Florianópolis-SC. The colors refer to periods of days where the incidence increases or decreases for at least three in a row. Only the upward phase is considered.

**Fig. 8.**
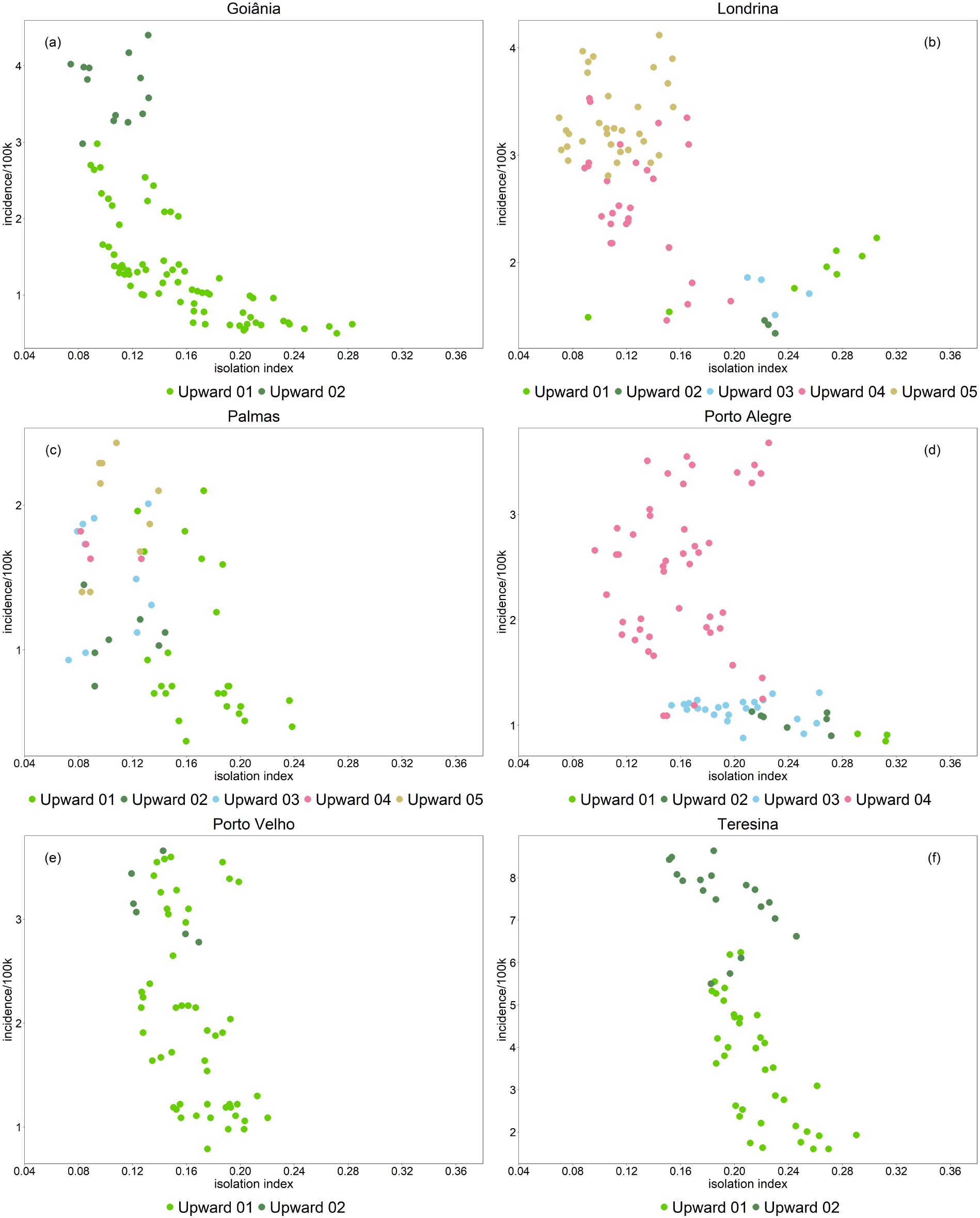
Dispersion between the daily incidence and the isolation index for the cities of ((a) Goiânia-GO, (b) Londrina-PR, (c) Palmas-TO, (d) Porto Alegre-RS, (e) Porto Velho-RO and (f) Teresina-PI. The colors refer to periods of days where the incidence increases or decreases for at least three in a row. Only the upward phase is considered.

The cities Teresina and Porto Velho, where the *L*-shape is not observed, are in the region of influence of Fortaleza and Manaus, respectively, Metropolises that did not implemented mitigation strategies capable of increasing significantly social isolation, what could have caused the mitigation on these cities to not properly function. Indeed, Teresina has sustained an increase on the incidence although having a high social isolation index during the whole upward phase.

Among the Metropolises, the number of monotone intervals varied between 2 and 5. Goiânia, the Metropolis in Midwest, had 2 intervals, Campinas, the Metropolis in the Southeast, had 3 intervals and the Metropolises in the South region had 3, 4 and 5 intervals (Curitiba, Porto Alegre and Florianópolis, respectively). The HDI values of these cities, in the order they were mentioned, are 0.799, 0.805, 0.823, 0.805 and 0.847. Those localized in the South region presented a greater number of intervals, indicating that the adopted measures in these cities succeeded on breaking the increasing sequence. In the case of Florianópolis, since a major part of its territory in an island, other geographical feature could have contributed to control the disease spreading.

For the Regional Capitals A, the numbers of intervals varied between 2 and 7. Teresina, localized in the Northeast region, had 2 intervals, while Cuiabá and Campo Grande, both in the Midwest region, had 5 and 7 intervals, respectively. The HDI of these cities, in the order they were mentioned, are 0.751, 0.785 and 0.784. In this hierarchy, there are only capitals from Northeast and Midwest regions and the latter ones presented better results.

Finally, for the 4 Regional Capitals B, the number of intervals varied between 2 and 5. Porto Velho, localized in the North region, had 2 intervals, while Palmas, from Midwest region and Londrina and Caxias do Sul, both from the South region, had 5 intervals. The HDI of these cities, in the order they were mentioned, are 0.736, 0.788, 0.778 and 0.782. The performance of Palmas on controlling the disease’s spreading could be explained by the influence of Goiânia over it. The Capital from the North region presented the worst result, that is, lower number of intervals, while Midwest and South regions presented the best results. This detailed analysis, considering the number of intervals, shows again the importance of the urban hierarchy, HDI and regional factors in the spreading of the disease.

Analyzing the results of each hierarchy, it seems that the number of intervals increases as the HDI increases (correlation coefficients 0.90, 0.98 and 0.76 for Regional Capitals A, Regional Capitals B and Metropolises, respectively). Although there seems to exists a threshold of the social isolation index below which an explosion of cases occurs, its value vary among the cities. Therefore, besides human mobility, other factors are certainly influencing the velocity of the disease’s spread.

## 4 Discussion

In this work, we explored some variables that can be responsible for the failure or success of the mitigation strategies to halt the spreading of the COVID-19 epidemic in the Brazilian territory. In particular, we focused on social isolation, since during this epidemic period it was one of the few strategies available, besides prophylactic measures such as the use of masks and personal hand-hygiene. Our analysis sought to associate two sets of temporal series, the social isolation index, and the incidence, exploring the relationship between these two data sets in different cities in Brazil looking for patterns and what can explain them.

The first thing that we could notice is that there is no direct and simple relationship between the social isolation index and the incidence. In fact, the evolution of the disease is driven by multiple factors. Besides the social isolation index, the urban hierarchy, the human development index and human and trade mobility together can explain the behavior of the epidemic, as shown in Sections 3.1-3.3. Among those patterns, we could find a relation between the starting time of the implementation of the mitigation strategy and the time at which the peak was reached. Namely, the cities that successfully implemented the social isolation strategy in stages 3 or 4 reached the peak of incidence later. Moreover, among those cities, a threshold on the social isolation index could be found. This threshold holds at the begging of the epidemic and promotes an efficient control of the incidence. With time, the social isolation index decreased, and the number of cases sharply increased in such a way that high isolation measures did not work anymore.

There are some limitations related to the data used here. The bias of the daily isolation index is not controlled. On top of that, the daily incidence data cannot capture the asymptomatic cases. Despite these limitations, we successfully pointed out some patterns related to these data. We also could hypothesize that at least two patterns of disease dispersion can be found in Brazil^29,35,28^: a hierarchical one at Southwest region (like São Paulo state)^15^ and a local diffusion type at Northeast (like Ceará state). We also hypothesize that the epidemic is synchronized in each Metropolis^17,39^. These are relevant topics to be pursued.

## Data Availability

Social distancing data, based on mobile geolocation, on city level for Brazil. Available at https://github.com/pedrospeixoto/mdyn/tree/master/data_mobility
All codes used in statistical analysis are available at
https://github.com/pedrospeixoto/mdyn

https://github.com/pedrospeixoto/mdyn

## Resource Availability

### Lead Contact

PSP.

### Data and Code Availability

The datasets and codes generated during this study are available at the GitHub repository Mdyn https://github.com/pedrospeixoto/mdyn.

## Acknowledgments

PSP was supported by grant # 16/18445-7, São Paulo Research Foundation (FAPESP) and by grant #301778/2017-5, National Council for Scientific and Technological Development (CNPq). The research of CPF is supported by grant #2019/22157-5, São Paulo Research Foundation (FAPESP) and by grant #302984/2020-8, National Council for Scientific and Technological Development (CNPq). DM was supported by the National Council for Scientific and Technological Development (CNPq) during the development of this paper.

## Author Contributions

Data Curation, Formal Analysis and Investigation were mainly performed by CPF, MPM and CMP. All other aspects of the paper had active participation of all authors, including conceptualization, methodology, discussions, writing and review.

## Footnotes

∗ Recently the company changed name to Incognia

† Due to a recent company shift in business area, the base suffered a reduction in size in 2021 and the company stopped providing the Social Isolation Index.

† Hence we excluded Jundiaí and São Paulo from this analysis.

